# Substance Use and other Correlates of HIV infection among transwomen and men who have sex with men in Perú: Implications for Targeted HIV Prevention Strategies for Transwomen

**DOI:** 10.1101/2022.08.27.22279296

**Authors:** Elena Cyrus, Javier R. Lama, Jorge Sanchez, Segundo Leon, Manuel V. Villaran, Panagiotis Vagenas, Daniell S. Sullivan, David Vu, Makella Coudray, Frederick L. Altice

**Affiliations:** University of Central Florida: Population Health Sciences, College of Medicine, Orlando, FL, USA; Yale School of Public Health, Department of Global Health, New Haven, CT, USA; Associación Civil Impacta Salud y Educación (IMPACTA), Lima, Perú; Centro de Investigaciones Tecnológicas, Biomédicas y Medioambientales; Universidad Nacional Mayor de San Marcos, Lima, Peru; San Juan Bautista Privada, Lima, Perú; Berkeley Research Development Office, University of California Berkeley, Berkeley, CA; Univeristy of Central Florida: College of Medicine, Orlando, FL, USA

**Keywords:** transwomen, MSM, HIV, risk behavior, substance use, Perú

## Abstract

**Introduction:** Characterization of HIV risk factors among transwomen and men who have sex with men (MSM) should be assessed separately and independently. However, due to several constraints, these populations continue to be conflated in clinical research and data. There are limited datasets globally powered to make such comparisons. The study aimed to use one of the largest surveys of transwomen and MSM in Latin America to determine differences in HIV risk and related correlates between the two populations.

**Methods:** Secondary data analysis was completed using a cross-sectional biobehavioral survey of 4413 MSM and 714 transwomen living in Perú. Chi Square analysis of selected HIV correlates was conducted to examine differences between transwomen and MSM. Additionally, stratified binary logistic regression was used to split data for further comparative analyses of correlates associated with transwomen and MSM separately.

**Results:** HIV prevalence among transwomen was two-fold greater than among MSM (14.9% vs. 7.0%, p<0.001). Transwomen had a higher prevalence of most HIV risk factors assessed, including presence of alcohol dependence (16.4% vs. 19.0%; p<.001) and drug use in the past 3 months (17.0% vs. 14.9%). MSM were more likely to use marijuana (68.0% vs. 50.0%, p<.001), and transwomen were more likely to engage in inhaled cocaine use (70.0% vs. 51.1%, p<.001). The regression exposed differences in correlates driving sub-epidemics in transwomen vs. MSM, with a trend of substance use increasing HIV risk for transwomen only.

**Conclusions:** Transwomen were more likely to be HIV-infected and had different risk factors from MSM. Targeted prevention strategies are needed for transwomen that are at highest risk. Additionally, further research is needed to determine if these observations in Perú regarding substance use patterns and the role of substance use in HIV risk relate to other trans populations globally.

## Introduction

Men who have sex with men (MSM) and transwomen in Perú bear a disproportionate burden of human immunodeficiency virus (HIV) infection.^1,2^ In 2016, the HIV seroprevalence among Peruvian MSM and transwomen in Lima was >10% and >20%, respectively, compared to the general Peruvian population prevalence of <1^%^.^3^ Similar to other Latin American countries, MSM and transwomen in Perú are socially and medically vulnerable communities, subject to discrimination, social exclusion and insufficient access to preventative and long-term care, partly because of cultural and sexual biases that are pervasive against same-sex relations in the wider Peruvian society.^4^ Additionally, MSM and transwomen are prone to engage in high-risk sexual behaviors, such as transactional or survival sex work and substance use.^2^

While acknowledging these common characteristics among MSM and transwomen, the scope of the problem and risk are not equal for both groups, and transwomen are at greater risk for HIV transmission.^2,3,5^ A combination of social, biological, and structural factors contribute to increased HIV vulnerability in transwomen. Deeply rooted stigma and discrimination in gender expression and gender identity can result in physical and psychological harm towards trans populations that can drive these individuals towards unhealthy coping mechanisms such as substance use.^6^ The lack of a legal identity status for transwomen also limits economic opportunities for these individuals, forcing them to pursue illegal and socially stigmatizing economic activities such as sex work, which could lead to increased risky sexual behaviors and subsequent HIV exposure.^7^ Furthermore, transwomen have been historically neglected in HIV research studies compared to MSM, contributing to the lack of preventative and treatment resources in these communities.^8^ Overall, these evidenced factors and more create increased HIV and substance use rates in transwomen populations compared to MSM and the general population.

Evidence of an enhanced disparity between MSM and transwomen can help guide policy or interventional studies that address the specific needs of transwomen. Due to limited resources and competing research priorities, or conflation for convenience, research endeavors often address these groups simultaneously.^9–11^ The purpose of this study was to further examine differences in these two groups from Perú’s largest biobehavioral surveillance study on correlates and sexual risk behavior among MSM and transwomen and to compare characteristics between them to justify the separation of these groups for singular and independent research efforts.

## Methods

### Study Design and Data Collection

Between May and October 2011, 5,575 Peruvian MSM and transwomen were recruited using modified snowball methods and peer-educator outreach in previously mapped venues frequented by MSM and transwomen in Lima and four other cities: Ica, Piura, Iquitos, and Pucallpa. Extensive details about recruitment and study procedures have been previously described.^1,2^ Eligibility included individuals 18 years of age or older, being genetically male, and self-reporting at least one male sexual partner in the previous 12 months. The two-step question was used to classify individuals as either transgender or MSM.^12^ Of the 5,755 MSM and transwomen recruited, 5,148 completed all study procedures and were eligible for analysis. Of these, 21 did not complete the sexual identity questions regarding gender identification and were thus excluded from the analysis, resulting in a final analytic sample of 5,127.

The research project was approved by the Institutional Review Boards of IMPACTA Perú and Yale University. After establishing eligibility, participants underwent informed consent procedures. Activities sequentially included pre-test HIV counseling, phlebotomy and urine testing, a computer self-assisted self-interview (CASI) for data collection, and post-test counseling and review of test results. Those testing positive for HIV or sexually transmitted infections (STIs) were treated according to Peruvian national guidelines.

### Variables

For analysis, participants’ ages were dichotomized as 25 years and younger or older than 25, based on median age. Residence was stratified as living within or outside of Lima. High-risk sexual behaviors associated with HIV infection were decided a priori using the Alaska sexual risk criteria^12^ for behaviors during the previous 6 months: 1) having an STI; 2) self-identification as a commercial sex worker; 3) no condom use during last anal intercourse; 4) anal intercourse with more than 5 partners; and 5) being a sexual partner of an HIV-infected male.

Sexual identity was based on whether participants self-identified as being MSM or a transwoman. Identification of transwomen was confirmed through a two-step question asking about gender assigned at birth and current self-identified gender.^13^ Commercial or transactional sex work (CSW) was defined as having exchanged sex for money, drugs or rent in the past 6 months. Standard Determine® (Third Generation) rapid enzyme-linked immunosorbent assay (ELISA) HIV testing provided preliminary positive HIV test results that were confirmed with Western Blot testing. Rapid plasma reagin (RPR) was used to detect the presence of syphilis and confirmed through microhemaglutination assay for Treponema pallidum antibodies (MHA-TP). Acute syphilis was based on having an RPR titer ≥1:16; this correlates with >90% likelihood of having an active infection.^4^ In addition to these two biomarkers to test for presence of HIV and syphilis, participants also reported any other STI diagnosis within the past 6 months. Additionally, we asked about the diagnosis of syphilis or any other STI in the past 12 months.

Alcohol use disorders (AUD) were assessed using the alcohol use disorders identification test (AUDIT), dichotomized by 8 or higher for hazardous use and 20 or higher for alcohol dependency.^14^ The AUDIT has adequate reliability and validity, and its sensitivity and specificity are comparable to those of other screening measures.^15^ Drug use was self-reported in the past 3 months and included any use of marijuana, ingested cocaine (often referred to as “pasta” in Perú) or inhaled/powdered cocaine, amphetamines, poppers, or ecstasy.

### Analysis

Statistical analysis compared HIV/STI prevalence, self-reported sexual risk behavior, and demographic characteristics between MSM and transwomen. All variables were binary or categorical, and categorical variables were ultimately dichotomized. Variables were first analyzed using χ^2^, and p-values<0.05 were considered significant. Multiple logistic regression was conducted separately comparing the correlates of HIV infection among MSM and transwomen. Parsimonious models where significant correlates on bivariate analysis at p<0.05 were included in the final model. Backward and forward elimination models were also conducted. Goodness-of-fit was determined using the Akaike Information Criterion, which was used to select the best fit model. Statistical Package for the Social Sciences (SPSS) 22 software was used for all analyses.^16^

## Results

Of the 5,127 participants, 714 (13.9%) self-reported as transwomen. Figure 1 shows comparisons of different characteristics and HIV-associated risk factors between MSM and transwomen, and Figure 2 illustrates the breakdown of illicit or illegal drug use by type of drug. HIV prevalence was significantly two-fold higher among transwomen compared to MSM (14.7% vs. 7.0%, p<.001) and for syphilis and acute syphilis (27.0% vs. 12.3%, p<.001; 10.8% vs. 4.2%, p<.001). In addition to the biomarkers, transwomen also had a higher self-report of syphilis or an STI in the past 12 months (13.0% vs. 6.3%, p<.001; 29.3% vs. 24.4%, p=.04).

**Figure 1:**
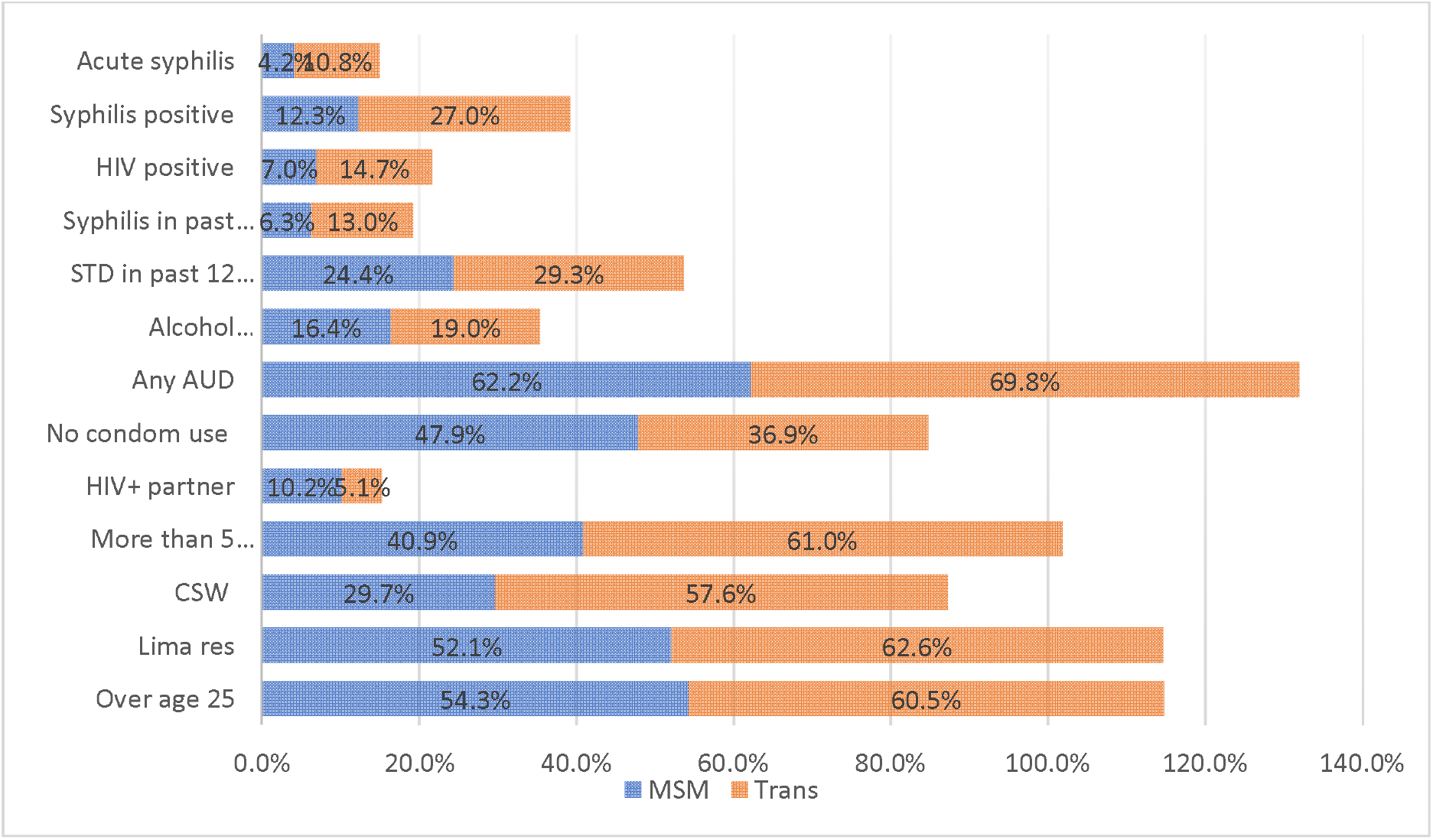
Demographics, risk factors, and STI/HIV prevalence by sexual identity: Transwomen versus Men Who Have Sex With Men.

**Figure 2:**
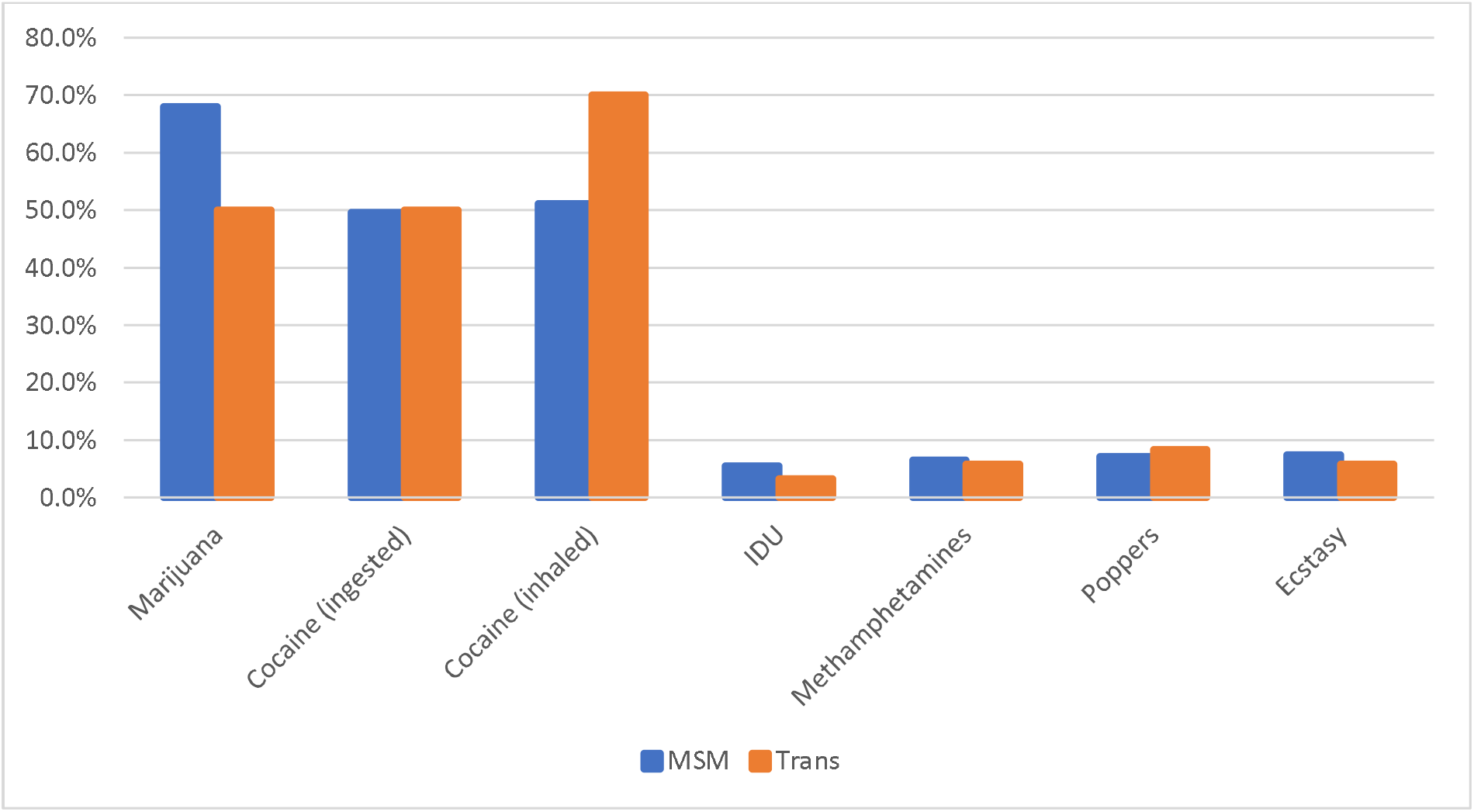
Illegal drug use by Transwomen versus Men who have sex with men (n=5,127)

Compared to MSM, transwomen were significantly more likely to have more than five sexual partners (61.0% vs. 40.9%, p<.001) and engage in commercial sex work (57.6% vs. 29.7%, <.001). However, more MSM reported engaging in unprotected sex in the last sexual encounter (47.9% vs. 36.9%, p<.001) and having an HIV infected sexual partner (10.2% vs. 5.1%, p<.001).

Compared to MSM, more transwomen met criteria for alcohol dependence (19.0% vs. 16.4%, p=.06). Differences in overall illicit/illegal drug use between transwomen and MSM were not significant (17.0% vs. 14.9%, p=.15; data not presented); however, in further analysis by drug type, there were significant differences in the two dominant drugs – inhaled cocaine and marijuana. MSM were more likely to use marijuana (68.0% vs. 50.0%, p<.001), and transwomen were more likely to engage in inhaled cocaine use (70.0% vs. 51.1%, p<.001).

For associations between HIV correlates and a positive HIV status, with the exception of alcohol dependence, the selected risk factors were all found to be significantly associated with MSM as depicted in Table 1. For transwomen, none of the correlates were statistically significantly associated at alpha ≤.05. However, at alpha ≤.10, living in Lima (OR=3.27, p<.001), having acute syphilis (aOR=1.81, p=.10), engaging in illicit/illegal drug use in the past three months (aOR=1.65, p=.08), and being alcohol dependent (aOR=1.69, p=.07) increased the likelihood of a positive HIV status for the study participant. Finally, being over the age of 25 was not significantly associated with HIV infection for transwomen, whereas in the MSM cohort, being over the age of 25 was associated with an increased risk of HIV infection.

**Table 1:** Independent Risks for HIV among Transwomen and Men Who Have Sex with Men (N=5,127)

| Variable | Men who have sex with men<br>(n=4413) |  |  | Transwomen (n=714) |  |  |
| --- | --- | --- | --- | --- | --- | --- |
|  | AOR | 95% CI | p-value | AOR | 95%CI | p-value |
| Over age 25 | 1.25 | .97 - 1.61 | 0.08 | 1.13 | .70 - 1.80 | 0.62 |
| Lima residence | **2.86** | **2.16 - 3.78** | **<.001** | **3.27** | **1.81 - 5.91** | **<.001** |
| Acute syphilis | **2.6** | **1.63 - 4.15** | **<.001** | 1.81 | .88 - 3.73 | 0.10 |
| Syphilis positive | **2.3** | **1.60 - 3.31** | **<.001** | 1.4 | .78 - 2.52 | 0.26 |
| Any STI past 12 mths | **2.02** | **1.50 - 2.71** | **<.001** | 0.96 | .51 - 1.82 | 0.90 |
| Drugs in past 3 mths | **0.49** | **.32 - .76** | **<.001** | 1.65 | .94 - 2.87 | 0.08 |
| Alcohol Dependent | 1.21 | .84 - 1.76 | 0.30 | 1.69 | .94 - 3.03 | 0.07 |

## Discussion

Study findings demonstrate transwomen living in Perú have a higher HIV/STI prevalence and engaged in more high-risk behavior than MSM. HIV prevalence among transwomen was more than double that of MSM, which is consistent with other research that identifies transwomen as a higher risk group.^17^ Transwomen were more likely to engage in sex work, to have a higher mean number of sexual partners, and to have an alcohol use disorder.

In this study, drug use in the past three months was statistically associated with HIV in both MSM and transwomen; however, it was protective for MSM, although statistically non-significant at the designated alpha level. Drug use also had a trend of increasing HIV risk for transwomen.

In-depth analysis was performed for drug use, and there were differences in the drug type usage – MSM had more marijuana use, while transwomen had higher rates for inhaled or snorted cocaine (Figure 2). Upon further exploration of the literature, marijuana, which in this study is more frequently used among MSM compared to transwomen, might be commonly used in a recreational capacity in MSM populations. However, cocaine use among transwomen may be used more often as a coping mechanism compared to MSM during transactional sexual encounters. ^18,19^ Theoretically, both scenarios of recreational marijuana use or cocaine during survival sex can increase sexual risk behavior and HIV risk.^12,18,19^ However, in this study, perhaps drug use, more specifically cocaine use, increases risk among transwomen because of greater exposure to and frequency of illicit/illegal poly substance drug use.

MSM were more likely to report having sex with an HIV-infected partner in the past three months than transwomen; however, transwomen also reported more commercial sex work and engaging in sexual activities with more than five partners. If transwomen engage in more commercial sex work and the HIV status of their sexual partner is not necessarily known, sexual acts with an HIV positive partner may be underreported among transwomen. In this population, little is known about the demography and health status of sex worker clients’; however, among those who engage in sex work, unprotected sexual acts with regular or non-paying clients are commonly reported^20–23^, and these unprotected sexual acts with regular clients can contribute to the spread of HIV/STIs.

HIV infection being associated with syphilis and older MSM is consistent with a previous similar study among Peruvian MSM.^24^ However, there are two differences between this study and the other study. First, age or being older than 25 does not seem to be related to HIV status for transwomen in the present study. Additionally, in this study, testing positive for syphilis was not associated with HIV infection among transwomen, despite syphilis prevalence being more than double among transwomen than MSM. Only acute syphilis or early infection, which was also more prevalent among transwomen, was associated with an HIV seropositive status. However, the lack of association between a non-reactive syphilis screening test and HIV seropositivity may be related to lack of power in the smaller sample size of the transwomen cohort.

The impact of having syphilis or co-infection with syphilis and HIV is problematic because syphilis can lead to a higher likelihood of HIV transmission, while being co-infected can accelerate progression of HIV illness.^25^ Therefore, this area deserves more investigation into possible confounders or moderators, such as access to care or other structural issues^26^, that impact the relationship between these two variables. A more stable statistical model should be used to confirm this relationship, especially now when there has been a further increase of syphilis rates in Perú and globally^27^.

Alcohol use and sex under the influence, which are known correlates for HIV transmission^12^, are more prevalent among transwomen than MSM living in Perú^28^. This statistic is consistent in this study. Furthermore, alcohol dependence is an area of concern for transwomen, as more transwomen were identified as being alcohol dependent in this study, and alcohol dependence was trending as being linked with positive HIV infection, which was not the case for MSM in the sample.

There were several limitations to this study. The method of recruitment, modified snowball sampling, may have introduced some bias; however, it is considered the most useful method to sample hard-to-reach populations such as MSM and transwomen.^29^ It should be noted that the sample may not be representative of the target population as compared to a typical random sample. The study was cross-sectional, which did not allow for a temporal assessment of causality. Recall bias may have also been an issue for some of the self-reported variables. With transwomen having one sixth of the MSM sample size and having a less than 15% HIV prevalence, the power may not have been adequate in the logistic regression analysis to clearly identify correlates associated with HIV-infection among transwomen. Although there were biomarkers for HIV and syphilis, self-report was used to capture data on other STIs and alcohol use, which could have led to some level of underreporting and underestimations.

## Conclusion

By using one of the largest samples of surveillance data among Peruvian MSM and transwomen, this study is one of few existing studies that examines differences in risk factors between MSM and transwomen. Our study provides evidence for the need of expanded and unique research efforts targeted towards transwomen in the Andean region and all South America. These results may also be replicated on a global scale. Treating these two groups as the same demographic may ultimately result in disappointing outcomes for primary prevention programs aimed at curbing HIV incidence among transwomen and ultimately a misapplication of valuable public health resources.

Both MSM and transwomen living in Perú engage in high-risk sexual behaviors, placing them at increased risk for HIV/STI transmission. However, transwomen have a formidable disadvantage between the two groups. Compared to MSM, transwomen engage in more high-risk behaviors such as substance and alcohol use, commercial or transactional sex-work, sex with multiple partners, and unprotected sex. Emphasis should be placed on this group as a most at-risk population (MARP)^28,30^ in Perú and globally.

Additionally, substance use, in particular alcohol dependence and inhaled cocaine use, may have an association with high-risk sexual behavior among transwomen, and this relationship should be explored further. Emerging research^5,6^ continues to reinforce the importance of these types of studies, as they are meaningful for developing a comprehensive body of literature on this subject and providing tools for HIV researchers working with transwomen.

## Data Availability

All data produced in the present study are available upon reasonable request to the authors.

## Author Contributions

Elena Cyrus, PhD, MPH^1, 2^: Principal Investigator of the study. I developed the research question, completed analysis and was the primary writer for the paper.

Javier R. Lama, MD, MPH^3^: Co-Investigator of parent study where the data was sourced in Perú. Contributed to review and edits of final version of paper

Jorge Sanchez, MD, MPH^4^: Co-Principal Investigator of parent study from where the data was sourced in Perú. Contributed to review and edits of final version of paper

Segundo Leon, MPH^5^: Contributed to interpretation of results and manuscript writing

Manuel V. Villaran, MD, MPH, MSc^3^: Contributed to interpretation of results and manuscript writing

Panagiotis Vagenas, PhD, MPH^6^: Contributed to interpretation of results and manuscript writing Daniell S. Sullivan^7^: Contributed to manuscript writing and editing

David Vu^7^: Contributed to manuscript writing and editing

Makella Coudray PhD^1^: Contributed to manuscript writing and editing

Frederick L. Altice, MD, MA^8^: Senior mentor/author and Co-Principal Investigator of parent study from where the data was sources

## Acknowledgements

This project was supported by the National Institute on Drug Abuse (NIDA) at the National Institutes of Health and Award K99/R00DA046311; The Loan Repayment Program at the National Institute on Alcohol Abuse and Alcoholism (NIAAA); and the NIH Research Training Grant # R25 TW009338 funded by the Fogarty International Center, the National Heart, Lung and Blood Institute, the NIH Office of the Director: Office of Research on Women’s Health and the NIH Office of the Director Office of AIDS Research.

